# Characterizing the leadership of family medicine registrars of Kamuzu University of Health Sciences: Perspectives of healthcare workers engaged in bedside teaching

**DOI:** 10.64898/2026.02.25.26347117

**Authors:** Tony Majo, Duncane kwaitana, Franchine Mambo, Martha Kabudula Makwero

## Abstract

Good leadership is a crucial aspect for a good primary healthcare system and for enhancing patient health outcomes. This qualitative exploratory study sought to explore the leadership role played by family medicine registrars in bedside teaching at Mangochi District Hospital and Nkhoma Mission Hospital in Malawi. Focus group discussions were held with healthcare workers who worked under the registrars, and the data was analyzed qualitatively using inductive and deductive analysis. From the study, it was established that the registrars have good clinical leadership skills, including working in collaboration, mentorship, ethical behavior, flexibility, and resourcefulness. However, the effectiveness of the leadership role is limited by a lack of clear role boundaries, mentorship, limited participation in system decision-making, and a lack of feedback. The family medicine registrars demonstrated high levels of interpersonal and professional competencies, which have a high potential for improving leadership roles. The study has provided useful insights on how the leadership training in the Master of Family Medicine course at Kamuzu University of Health Sciences (KUHeS) can be improved.

## Introduction

The Astana Declaration (2018) reaffirmed the commitment of global leaders, first expressed in the Alma-Ata Declaration (1978), to pursue Health for All through strong, people-centered primary health care (PHC) systems. The declaration emphasized making PHC accessible, equitable, safe, high quality, comprehensive, efficient, acceptable, available, affordable, and gender-sensitive (1). Despite this commitment, several factors such as lack of sustainable political will, weak monitoring and evaluation systems, and unsustainable financial models, continue to threaten the attainment of universal health coverage (2).

In response, waves of reforms, including strengthening governance and leadership, have been introduced globally (3). Strong and effective leadership has been identified as a critical driver of health system performance and improved health outcomes (4). Evidence indicates that good governance and leadership at all levels of the PHC system are essential to ensure that the needs of patients, clients, and healthcare providers remain central to health sector priorities (5).

Equipping leaders within PHC systems with the necessary leadership skills is therefore imperative for inspiring and managing healthcare workers to achieve organizational goals (3). The World Health Organization (WHO) has endorsed leadership training as a key health systems strengthening strategy. In sub-Saharan Africa, Leadership Development Programs (LDPs) and leadership-focused training have increasingly been integrated into pre-service curricula and in-service training to build the capacity of healthcare managers (4).

In Malawi, the Health Sector Strategic Plan III (HSSP III) highlights leadership and governance as priority areas for reform, recognizing persistent challenges such as:

- Weak leadership capacity to plan, execute, and evaluate policies and plans.
- Ineffective enforcement and monitoring of health sector policies, legal, and regulatory frameworks at national and sub-national levels.
- Weak accountability mechanisms and performance management at service provider level (6).

These gaps mirror findings that leadership capacity, particularly at district and facility levels of Malawi’s PHC system, is insufficient to cultivate a performance culture aligned with PHC goals and priorities (5). Against this backdrop, leadership training for family medicine registrars introduced at KUHeS, is a critical intervention. Such training provides future healthcare leaders with the competencies needed to strengthen PHC delivery and governance in Malawi. Since the introduction of the program in 2020, there has been no study aimed at characterizing the leadership abilities of FM registrars. This study therefore sought to characterized the leadership of family medicine registrars during bed – side teaching through the perspectives of the supervisees.

## Methods

### Study Design

This was an exploratory descriptive qualitative study using semi-structured focus group interviews.

### Study setting

The study was conducted at Mangochi district and Nkhoma mission hospitals which offer bed – side teaching under family medicine masters program hosted at KUHeS. Kamuzu University of Health Sciences is one of the public universities in Malawi which was established in 2019 following the merger of the Kamuzu College of Nursing and the Malawi College of Medicine. Guided by its mission to deliver high-quality education, research, and healthcare services, KUHeS plays a central role in advancing Malawi’s health sector. The university offers a broad range of undergraduate and postgraduate programs designed to prepare competent health professionals and leaders. Among its flagship programs is the Bachelor of Medicine, Bachelor of Surgery (MBBS), alongside specialized training in nursing, public health, pharmacy, biomedical sciences, and various clinical disciplines.

In 2020, the Department of Family Medicine (FM) introduced a two-week leadership training program for third year registrars in Master of Medicine (MMed) in Family Medicine. These registrars are tasked with significant responsibilities in supervising healthcare workers during bedside teaching. Their supervisory roles demand strong leadership abilities, including the capacity to create a positive learning environment, motivate supervisees, and drive productivity within healthcare teams. The leadership training program was therefore developed to equip registrars with the knowledge, skills, and attitudes necessary to: 1) Manage healthcare teams and resources effectively 2) Inspire and motivate colleagues and learners 3) Lead innovations and organizational change 4) Contribute to making health systems more resilient to external shocks. The program is structured around five core modules, reflecting essential domains of healthcare leadership:

1. Leadership for Self – personal leadership and self-awareness.
2. Engaging Others – teamwork, collaboration, and communication.
3. Achieving Results – goal-setting, accountability, and performance management.
4. Leading Change – driving and sustaining innovations in healthcare.
5. Management – practical aspects of planning, resource use, and decision-making.

Delivery of the modules is facilitated by KUHeS lecturers with formal qualifications in leadership and management. Following the training, registrars are deployed to supervise the six-week family medicine rotations in Mangochi District Hospital, and Nkhoma Mission Hospital. We purposively targeted Nkhoma Mission Hospital and Mangochi District Hospital.

### Theoretical Framework

The study was framed within the Six–Part Model for Adapting and Thriving during Transformation Change, serving as reference for the development of data collection tools and interpretation of results. This theory, provides a direction for health systems to adapt and thrive during period of transformation change (7). The five domains for the leadership training program align well with the six-part model for change. For example, self-awareness enables leaders to recognize the need for change, while teamwork and communication help create and share a clear vision. Goal-setting and accountability drive measurable progress and short-term wins, and leading change ensures innovations are implemented and sustained. Practical management skills in planning and resource use support all stages, making the change achievable and lasting. Together, these domains provide the essential capabilities for guiding and embedding effective change in healthcare systems. The model outlines six interrelated pillars as defined in Table 1. The pillars are considered essential for success in navigating transformative change (Figure 1).

**Figure 1.**
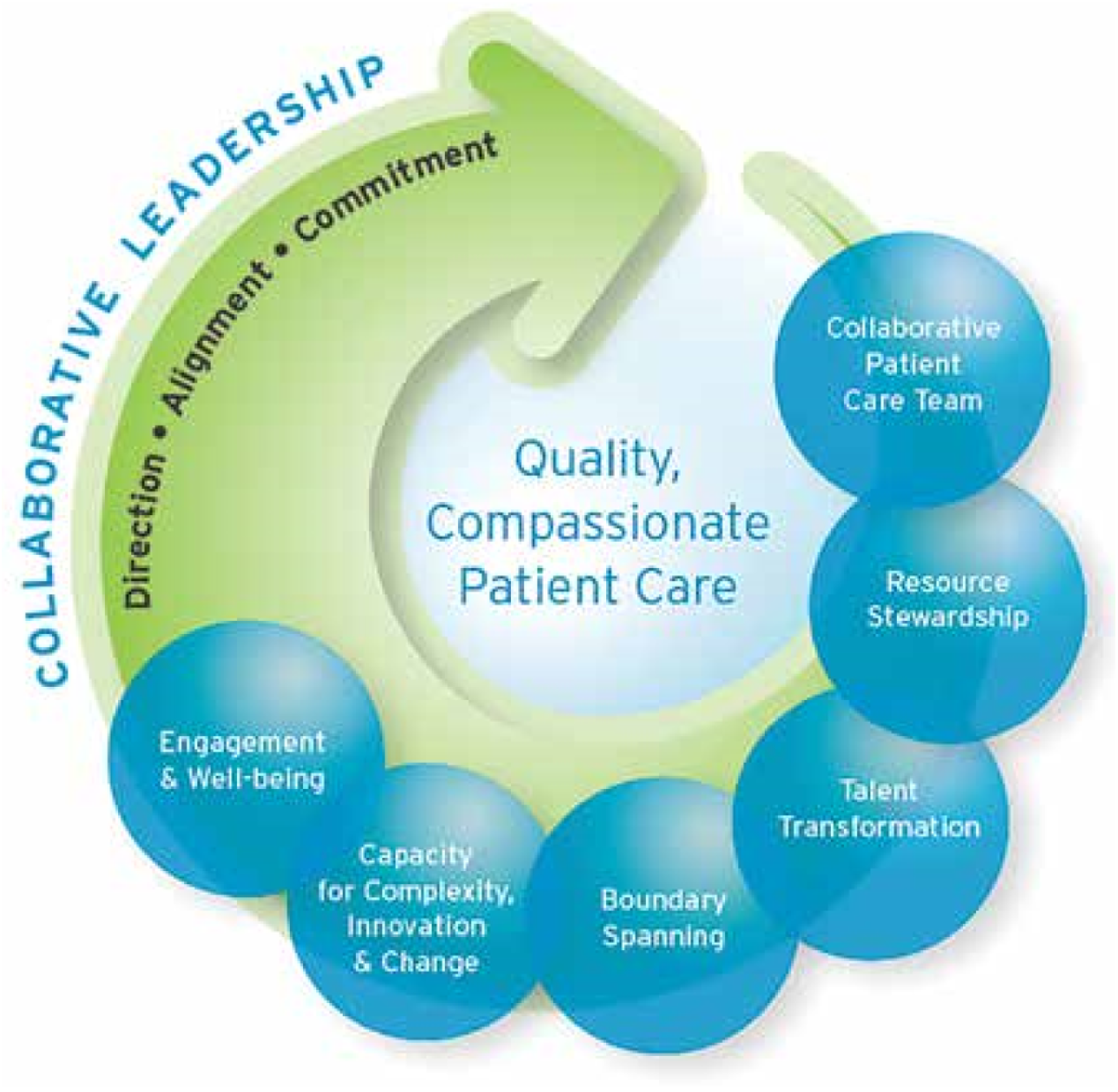
Six–Part Model for Adapting and Thriving during Transformative Change *Source: Browning, Torain & Petterson (2016, p.1)* (7)

**Table 1:** Six – Part Model pillars and definitions.

| **Pillars** | **Definition** |
| --- | --- |
| **Collaborative patient–care teams** | This pillar highlights the need for effective collaboration in interdependent patient-care teams |
| **Resource stewardship** | In resource-limited settings, leaders must use resources wisely while ensuring high-quality, compassionate care. |
| **Talent transformation** | Transformational leadership inspires and develops teams, fostering strong relationships, engagement, innovation, and accountability |
| **Boundary spanning** | Healthcare leaders must transcend traditional boundaries to enable collaboration across silos and support integrated, system-wide improvements |
| **Capacity for complexity, innovation, and change** | Health systems are increasingly complex, and leaders must anticipate and adapt to changes while fostering innovation and resilience |
| **Employee engagement and well-being** | Sustainable healthcare relies on engaged, healthy staff; leaders should support well-being and align employees with organizational goals |

*Adapted in Browning, Torain & Petterson (2016, p.1)*

### Study Population and Sample Size

The study focused exclusively on supervisees engaged in bed-side teaching, which runs for six weeks. Bedside teaching has long been regarded as a core instructional approach through which the essential components of clinical practice can effectively demonstrated and taught. It reinforces classroom learning by allowing mentors or supervisors to model key clinical skills, attitudes, and communication practices in real patient – care settings, while providing an opportunity to observe the mentee’s or supervisee’s competencies (8). Participants picked were those in their fifth or sixth week of training, as they are expected to have acquired sufficient experience to provide well-informed reflections on leadership during the teaching period. During bedside teaching, family medicine registrars supervise various healthcare workers (HCWs), including student nurses, student clinical officers, qualified nurses, qualified clinical officers, interns, and Bachelor of Medicine, Bachelor of Surgery (MBBS) students. This study purposively targeted Nkhoma Mission Hospital and Mangochi District Hospital as these are the only sites which offer bed – side teaching, under the supervision of the FM registrars. From each site, we conveniently selected one supervisee from each category of HCWs. However, MBBS students were not included, as none were available at the time of data collection, contrary to initial expectations. To address this, we conveniently picked a sixth participant from the available categories of HCWs. Consequently, a qualified clinical officer in Nkhoma and a nurse in Mangochi as the sixth participant in each study site, yielding a total of 12 participants across the two-study sites.

The sample size was guided by the principle of information power (9), which posits that the more relevant and information-rich the data, the smaller the required sample. This is consistent with Lincoln and Guba (1985), who recommend determining sample size based on information redundancy, whereby sampling is terminated once no new insights are generated. A sample size of 4 to 10 has been recommended by some authors as adequate for achieving data saturation in qualitative exploratory case study designs (10). In this study, data saturation was reached with the selected sample size, as the discussions yielded no new information or perspectives by their conclusion.

### Data Collection Tools and Procedures

The study received ethical approval from College of Medicine Research Ethics Committee the before the commencement of data collection. Formal permission was obtained from authorities at both study sites, and potential participants were notified in advance. Individuals were approached privately and provided with detailed information about the purpose of the study, data collection procedures, and their rights to decline or withdraw at any stage. Semi-structured interview guides were developed based on the Six–Part Model for Adapting and Thriving During Transformational Change, which provided the theoretical framework for examining leadership practices within the six-week rotational program.

Participants’ sociodemographic characteristics including age, sex, years of experience, and educational level were recorded; however, years of experience and professional qualifications were excluded for student participants, as these variables were not applicable (see Table 1). To ensure data quality and consistency, two trained research assistants, fresh graduates with bachelor’s degrees from public universities in Malawi conducted the focus group discussions. They were selected for their academic qualifications and independence from the study topic, having no prior involvement in leadership training or connections to participants, thereby minimizing potential bias.

A pilot study was conducted with two clinical officers through in-depth interviews. These officers had previously experienced bedside teaching under Family Medicine registrars and were pursuing further training at KUHeS at the time of the study. Insights from the pilot revealed that certain elements of the Six–Part Model were insufficiently covered in the initial interview guide. As a result, the tool was refined by adding some questions, in order to align it well with the framework to ensure it is contextually relevance to key leadership components, including collaboration, resource use, talent development, boundary spanning, adaptability to complexity, and employee engagement. Focus group discussions were conducted in private and convenient hospital locations to maintain comfort and confidentiality. With participants’ consent, all sessions were audio-recorded, and each discussion lasted approximately 1 hour and 20 minutes. All individuals who were approached agreed to participate, and no one declined.

## Data Analysis

The study adopted five-step process to analyze the qualitative data, which is consistent with Braun and Clarke’s thematic analysis framework (11). The steps included transcription, familiarization, coding, categorization, theme development, and interpretation. Data were analyzed using both deductive and inductive approaches. The deductive component was informed by the study objectives, while the inductive component allowed themes to emerge directly from participants’ narratives. This combined approach ensured a comprehensive and rigorous analysis of the data.

### Trustworthiness

To enhance trustworthiness of the findings, we employed Lincoln and Guba’s trustworthiness criteria (12). First, peer debriefing was undertaken by presenting the preliminary findings to an independent qualitative researcher from the field of education. This process allowed for an external examination of the methodological decisions, emerging results, and interpretive claims, thereby providing constructive critique and strengthening the rigor of the analysis. Second, member checking was conducted by re-engaging participants to verify the accuracy, resonance, and completeness of the study’s interpretations. These strategies collectively contributed to ensuring the credibility and trustworthiness of the findings

The study team comprised a medical professional, who holds a PhD, has extensive experience in primary healthcare research, and has conducted numerous qualitative studies in Malawi. This member also teaches at the postgraduate level. The team further included an education specialist in leadership and management with postgraduate qualifications and over ten years of experience in primary healthcare qualitative research. Their collective role was to provide technical oversight and guidance to the broader team.

### Ethical considerations

Ethical clearance to conduct this study was obtained from the College of Medicine Research and Ethics Committee (COMREC) (No. P.01/25 -1372). The study was also approved by the DHOs of the participating districts. Further, we sought written consent from the participants for being interviewees and allowing the interviews to be audio - recorded. We adhered to the Declaration of Helsinki’s ethical principles, safeguarding the rights and welfare of human participants in this study.

**Table 1:** Participants’ social demographic characteristics.

| **Categories of respondents** | **Mangochi**<br>**n = 6** | **Nkhoma**<br>**n = 6** | **Total**<br>**n = 12** |
| --- | --- | --- | --- |
| Student nurses | 1 | 1 | 2 |
| Student medical officers | 1 | 1 | 2 |
| Qualified nurses | 2 | 1 | 3 |
| Qualified medical officers | 1 | 2 | 3 |
| Interns | 1 | 1 | 2 |
| Age in years (average) | 29.5 | 25.5 | 27.5 |
| Sex |  |  |  |
| Males | 04 | 05 | 09 |
| Female | 02 | 01 | 03 |
| Years of experience<br>(excluding students) | 3.4 | 2.0 | 2.7 |
| Level of education<br>(excluding students) |  |  |  |
| Certificate | - |  |  |
| Diploma | 03 | 03 | 06 |
| Degree | 01 | 01 | 02 |

### Findings

The deductive analysis, guided by the Six–Part Model for Adapting and Thriving During Transformational Change, revealed key themes aligned with the model, including relationship building, innovation culture, resource management, and boundary spanning. Complementing these pre – determined themes, inductive analysis uncovered emergent pattern, notably ambiguity surrounding hierarchy, roles and responsibility and clinical care practices.

### Ambiguity on hierarchy, roles and responsibility

The majority of respondents reported limited awareness of the specific roles of registrars during Family Medicine (FM) bedside teaching. This gap in understanding was largely attributed to registrars not clearly communicating their responsibilities as bedside supervisors to the supervisees. Participants noted that the absence of explicit explanations regarding the registrars’ teaching and leadership duties created uncertainty about expectations and responsibilities. The majority of participants reported being unfamiliar with the role of Family Medicine registrars. Many noted that registrars did not make sufficient efforts to orient supervisees on their bedside teaching responsibilities, thereby leaving supervisees to rely on assumptions and guessing regarding their roles. Some participants further suggested that registrars themselves appeared uncertain about their supervisory functions, which consequently impacted their self – esteem and confidence in discharging their duties as bed – side supervisors.

> *"For a registrar to be recognized, they need to make efforts to know the staff on the ground, rather than just being seen as new people who go straight to the supervisor. Before they come here, they should already know their roles, especially as supervisors. With that awareness, they can build confidence and self-awareness, so that no one overshadows what they are doing. They should know their place, work hand in hand with us, and follow their objectives so that the project has a long-term impact.”* **(Participant 1).**

Furthermore, many supervisees reported being confused regarding the appropriate reporting lines within the clinical hierarchy. They were often unsure whether to direct questions, feedback, or clinical updates to the registrar or to the supervising consultant, which occasionally led to miscommunication and delays in clinical decision-making.

In addition, several participants indicated that the presence of supervisors sometimes overshadowed the registrars, reducing their visibility and authority in clinical and teaching settings. This overshadowing was perceived to limit opportunities for registrars to exercise leadership, mentor supervisees effectively, and develop confidence in managing complex clinical situations.

> *"In my case, I sometimes see more action from the supervisors than from the registrars themselves. Many people assume the supervisor will do most of the work, but I am able to differentiate between who is the supervisor and who is the registrar in question. Even in interviews with registrars, it has been noted that their input can seem minimal compared to when they work in a more informal setting, because the supervisor tends to overshadow them. That’s why I think there should be a clearer difference in roles between the supervisor and the registrar."***(Participant 5)**.

The combination of unclear roles and hierarchical ambiguity was therefore seen as a barrier to both registrar leadership development and optimal supervisee engagement during bedside teaching. Majority of participants observed that lack of role clarity has drained the confidence of registrars who seem to be fearful in the presence of their supervisors:

> *“Even in situations where we would expect registrars to take the lead—like cases that are beyond an intern’s level—they sometimes step back and say, ‘I can’t manage this, I need my supervisor.’ Yet, as people who have already finished their degree and started their master’s, we would expect them to have more confidence and an upper hand compared to interns.”* **(Participant 5)**.

> *“From what I observe, the loudest voice is always with the supervisor, and the registrar gets overshadowed. For example, when taking a patient to theater, the registrar often waits for the supervisor to say, ‘go ahead.’ When the supervisor is present, the registrar seems to hold back, as if they cannot fully function on their own. This makes it difficult for their light to shine”* **(participant 2)**.

### Relationship Building

The majority of respondents emphasized that cultivating strong relationships among supervisees, and between supervisees and their supervisors, is a critical determinant of organizational performance and team effectiveness. Participants, though mentioning that registrars have not been very dominant in their role as bed – side supervisors, reported that interactions within the supervisee group and with supervisors were largely cordial, supportive, and characterized by mutual respect likened it to a father – son dynamic.

> *I would describe my relationship with the supervisor as a ‘father and son’ relationship. From the first day in the medical ward, the supervisor’s attitude made me feel very comfortable. I can express myself without fear, and together we help patients using our combined knowledge. The supervisor, along with the nurse training officers (NTOs), teaches me on a day-to-day basis, which makes the relationship supportive and nurturing rather than strict or intimidating* **(participant 7)**.

These positive relationships were identified as key sources of encouragement, motivation, and professional growth, enabling supervisees to learn collaboratively, consult on clinical and educational matters, delegate responsibilities efficiently, and share challenges openly without fear of judgment:

> *Our supervisors [registrars] are also on the ground working with us, so things usually go as planned instead of just leaving tasks for the juniors. They even ask for our suggestions on what to do with a patient and appreciate it when we come up with good ideas. They inspire us a lot and I’ve had a very good interactions with the registrars and other healthcare staff, and overall, the relationship has been excellent* **(participant 11)**.

While respondents acknowledged the strength of these relationships, many were unable to articulate specific actions or strategies employed by registrars to establish or sustain this positive dynamic. Despite this, they consistently highlighted that certain cultural attributes such as open information sharing, adaptability, flexibility in task management, and a commitment to continuous learning which have built trust, have played a central role in consolidating trust and cohesion within the team.

> *"To me, I feel like most of the time it’s trust that makes a relationship. Because, at first sight, whenever we meet new people and they introduce themselves to us with their titles, we put our trust in them. We believe that with the purpose they are here for, and with the title they carry, this is what we are going to get from them, and this is the kind of direction we will be receiving. So even from the first point of contact, when new people join our wards, we have that trust and we say, I think now we are going to start making decisions at another level. From there, the relationship grows, because we know for sure that if things become tougher for us, we have this person to consult. And the more we consult them, the more positive feedback we get, and the more we are able to connect."* **(participant 8)**.

### Innovation Culture

Innovation is widely recognized as a cornerstone of organizational growth, positioning institutions to remain resilient and adaptable in the face of external shocks such as emerging disease outbreaks or shifting healthcare demands. Despite this, most respondents struggled to clearly articulate the nature of the innovation culture within their organization. When asked about existing strategies or frameworks that promote innovation, the majority were unable to identify specific initiatives, suggesting limited awareness or visibility of organizational efforts in this area.

> *Over the past three months, I haven’t witnessed any discussions with registrars about innovations within the organization. While during their training, supervisors challenge them to identify gaps and suggest improvements, so far, I haven’t seen any of these ideas put into practice**."*****(participant 1).**

Interestingly, when probed further, respondents generally described the environment as favorable for innovation in theory, but lacking in practical mechanisms that actively encourage creativity or risk-taking. No participant was able to cite a concrete example of innovative ideas generated by supervisees, nor instances where innovation was formally recognized or celebrated. This absence of visible innovation practices or success stories appears to show that culture of creativity remains underutilized. For example, a good portion of respondents noted that the organization demonstrates a markedly cautious approach towards new ideas, typically demanding robust evidence of assured positive outcomes prior to piloting or implementation, particularly when such innovations are proposed by junior staff.

> *The system at [name withheld] is flexible, but sometimes it depends on who brings an idea. A suggestion from someone junior might not be accepted as easily as one from a senior person, even if it is good. However, if the idea is evidence-based and well defended, it can be adopted regardless of who presents it. Medicine relies on proof, and when you can demonstrate that something works, the team is more likely to accept it. I haven’t personally seen it often, but there have been situations in the ward where evidence-based ideas were successfully introduced with a registrar’s support,* **(participant 11)**.

Overall, the findings reveal an organizational paradox: while the environment is not overtly resistant to innovation, it equally lacks the intentional strategies, flexible structures, and cultural reinforcements necessary to foster and sustain it. For example, when participants were probed about key characteristics of an innovative organization, such as a culture of experimentation, professional autonomy, risk-taking, employee recognition, dedicated budgets for innovation, and motivation for innovators, none were able to associate these features with their current organizational context.

### Resource Management

Participants’ feedback on resource management underscored the system’s commendable efficiency and effectiveness in the utilization of human, financial, material, and time resources. Within the broader theme of resource management, attention was directed to two selected domains. First, employee management, with particular emphasis on strategies for developing healthcare workers’ knowledge and skills and fostering motivation to enhance performance and resource mobilization. Second, material management, focusing on the availability and utilization of ward-based resources such as drugs and equipment during bedside teaching. These domains were identified as critical entry points for understanding how resources are managed in this setting. Material management was viewed by majority of participants positively, indicating that the system minimizes wastage through careful allocation, structured tracking, and accountability checks during handovers.

> *“In our ward, when medication is dispensed, it is given directly to the patient, who keeps it safely. The system ensures accountability, balances are tracked, records show who received what. This makes it difficult for anyone, even a student, to take medication improperly, because if it goes missing, the responsible person is exposed. Ultimately, patients and their guardians collect and safeguard their medication, ensuring proper use and security.”* **(participant 1)**

> *“During board or departmental meetings, ideas on how to use resources efficiently are regularly discussed. For example, when I was in the medical ward, the registrar often raised questions about how to implement best practices and improve processes, including deflection of diagnosis and other resource-related decisions within the ward”* **(participant 7)**.

This level of stewardship not only safeguards scarce resources but also reinforces a sense of collective responsibility among staff. In relation to human resource management, participants emphasized the supportive environment fostered through regular feedback sessions in the wards. These sessions were praised for promoting reflective practice, enhancing clinical competence, and contributing to professional growth.

> *I would say we get feedback multiple times. For example, in February, I received about four positive comments. Sometimes the feedback is simply acknowledging that what you did was good, without adding anything further. Other times, they might suggest minor improvements, but overall, the feedback is encouraging.* **(Participant 3**).

However, two areas were consistently flagged as requiring attention to fully optimize the system. First, participants noted the absence of a structured performance measurement framework to systematically assess, rank, and recognize supervisees’ exceptional contributions.

> *Usually, feedback happens within the ward, especially for students. However, we don’t have a formal system for appraisal by the registrar. The wards operate in teams, and immediate supervisors within those teams usually provide appraisals. Family Medicine personnel also have their own supervisors who appraise them depending on where they are working. In the labor ward, for example, we function as a team, so there isn’t always a clear system where one person oversees another specifically for appraisals* **(participant 11)**.

Furthermore, the majority of participants observed that there had been no deliberate initiatives aimed at mobilizing resources beyond the existing pool. This limitation was seen as a barrier to sustainability and innovation, as reliance on the same constrained resources restricted opportunities for growth, flexibility, and improved service delivery.

### Clinical care practices

When asked about situations where healthcare workers (HCWs) were required to respond to challenges beyond routine clinical care, most respondents were silent and did not cite specific experiences requiring supervisory intervention. This apparent silence could suggest that such cases are either rare, underreported, or perceived as part of routine professional judgment rather than extraordinary challenges.

Nevertheless, a few illustrative examples were shared. One respondent recounted a critical incident involving an accident victim who categorically refused a blood transfusion due to religious beliefs, despite the clinical team’s repeated efforts to emphasize its necessity for survival. The case was reported to the supervisor for further guidance, yet the team ultimately respected the patient’s autonomy, adhering to ethical principles of informed consent and non-maleficence.

> *“we faced this challenge where a woman with severe bleeding after delivery. She had lost so much blood, and we recommended a transfusion, but she was a Jehovah’s Witness and refused. Even after the registrar tried to explain the seriousness, she still declined, and unfortunately, she died. We later discussed it during ODD, and while some factors came from the referring hospital, ultimately it was also the patient’s refusal that contributed to the outcome. In such situations, it’s very difficult, because even when you know the right procedure, you cannot always apply it* **(participant 8)**

This situation highlights how HCWs must often navigate the delicate boundary between advocating for life-saving interventions and respecting individual rights and cultural values. Another participant praised the registrar’s initiative in responding to a different challenge, supporting a critically ill patient who could not afford essential medication. In this case, the registrar intervened to ensure the patient accessed treatment, demonstrating a commitment to equity and patient-centered care that extended beyond standard procedural expectations.

> *Yes, there have been situations where we faced critical decisions. For instance, we had a patient who had already received about 50 doses of atropine but still needed more. The question of why so much medication was being administered arose. In that moment, the registrar present advised that we should continue managing the patient according to their needs and only discuss further actions once the patient was stable. The patient eventually recovered, and this scenario highlighted the tension between protocol, oversight, and immediate clinical judgment.* **(participant 11)**.

Taken together, these examples demonstrate that while HCWs rarely reported challenges that forced them to cross professional or ethical boundaries, registrars were occasionally called upon to exercise judgment in ethically complex and socially sensitive situations. Such boundary-spanning actions reflect emerging leadership skills, particularly in balancing professional standards with patient autonomy, resource limitations, and the broader social determinants of health.

### Challenges and Recommendations

A recurring concern raised by participants was the need for registrars to embrace a stronger orientation culture, particularly in clarifying their leadership roles and responsibilities. Many supervisees observed that the boundaries between registrars and their supervisors were often blurred, resulting in uncertainty about reporting structures. This lack of clarity sometimes left supervisees confused about whom to approach for guidance, feedback, or problem-solving.

> *When a Family Medicine registrar joins the team, they usually introduce themselves and start working on the ground. However, it often takes time for the team to understand their exact role. Many just see them as another doctor, without knowing what is expected. If the registrar’s responsibilities were clearly communicated to the team from the beginning, it would help everyone understand their role and make the integration smoother."***(Participant 1)**.

Such ambiguity, if unaddressed, risks undermining both efficiency and accountability within the team. Participants therefore recommended that registrars take a more proactive role in outlining expectations, delineating responsibilities, and communicating clearly with both supervisors and supervisees.

Beyond role clarity, participants stressed the importance of registrars investing in relationship-building with staff at all levels of the health system. They emphasized that registrars should not only lead from a position of authority but also make deliberate efforts to know the staff personally, engage with them during ward rounds, and create a sense of partnership in service delivery. This approach would help registrars earn trust, improve team cohesion, and enhance the visibility of their leadership role. For supervisees in particular, such engagement would reduce hesitation in approaching registrars, thereby creating a more supportive and empowering learning environment.

> *"When a registrar is assigned to a ward, like the antenatal ward, they take on that leading role. They handle the ward round and make sure all the cases are attended to, while also helping the rest of us to finish everything else. But sometimes, they may not be available the whole day— maybe in the morning they are with us, and in the afternoon, they have other responsibilities. In those cases, we just accept it and continue working on our own. When they are there, they are our friends who help us, and when they are not, we function alone. I think it would also help if ward staff were oriented about the role of registrars, because some people understand what they are here for, but others don’t. Orientation would give everyone knowledge of what registrars are supposed to do."* **(participant 12).**

Participants also noted that registrars, as emerging leaders, should demonstrate greater adaptability in working hand-in-hand with both clinical and non-clinical staff. This involves not only supervising bedside teaching but also understanding the broader operational realities such as resource constraints, patient flow, and staff dynamics that shape day-to-day service delivery. By aligning themselves more closely with the ground realities, registrars would be better positioned to motivate supervisees, model problem-solving skills, and foster a culture of teamwork.

Overall, the recommendations call for registrars to embrace a leadership style characterized by clarity, visibility, and collaboration. Building an orientation culture, clarifying boundaries, and cultivating authentic relationships with staff were all seen as critical steps in strengthening their leadership presence. Embedding these practices into the leadership training program could ensure that registrars not only acquire technical knowledge but also develop the relational and organizational skills needed to lead effectively in complex healthcare settings.

> *“At this period, we just know it’s a rotation, but we don’t know the objectives. What we know is merely that they are just going to be seeing patients, basically. Maybe the problem is from the head of department, because when registrars are sent to our ward there should be proper coordination—like saying, this registrar is going to do this and that. But instead, when they are introduced, they just say, ‘we have a new visitor by such a name from family medicine,’ and stop there. So, the challenge is lack of proper communication from supervisors, registrars themselves, or ward in-charges about what exactly their objectives are. In fact, many health workers do not really understand what family medicine people are supposed to be doing, and if something could be developed to create awareness, others would better understand their roles and leadership responsibilities”* **(Participant 1).**

### Discussion of findings

This study characterizes the leadership of FM registrars during bed – side teaching through the perspectives of supervisees. The findings show that registrars play an important role in leading day-to-day clinical work and supporting the learning of junior staff. However, their leadership skills were stronger in some areas and weaker in others. These results seem to echo the findings showing that young medical managers often take on leadership roles while they are still learning how to lead effectively (13,14). One strong area of leadership they demonstrated was teamwork and collaboration. This kind of leadership is known to build trust and improve how teams work together (15–17). Teamwork is also considered crucial for establishing effective roles and achieving desired results particularly in service organizations (16). However, the team characterized during the bed -side teaching mainly happened within the ward. Registrars had little involvement in bigger, hospital-level decisions, which could be a sign that they are still developing system-level leadership skills (18). Given that roles of patient care teams are highly interdependent in nature, collaboration skills are hence crucial. The Six – Part Model for Adapting and Thriving during Transformation Change recommends the need to embrace collaboration skills beyond the workplace. To achieve this for instance, there is need to among others develop a learning culture within and outside the work environment as a platform for information sharing and testing (19). Another area was resource management. Registrars were careful with time, equipment, and other supplies. This is important in low–resource setting like Malawi where resources are limited (20). However, registrars mainly focused on using resources correctly, and they were less involved in finding new resources or advocating for more support. Health leadership frameworks say leaders should balance efficiency with innovation and problem-solving (7). Efficiency and effective management of resources is viewed as not only crucial, but also obligatory amidst increasing demand on accountability (21,22). The Six – Part Model for Adapting and Thriving during Transformation Change proposes the need for healthcare leaders to find means and ways of not only reducing costs, but also enhancement of investments and partnerships (7). This notion demands that leaders should develop the sense of ownership of the challenges facing the system (23). Registrars, also played an important role in teaching and mentoring. Many supervisees said registrars guided them, gave feedback, and acted as role models. This is consistent with leadership theories that show that good relationships between supervisors and supervisees improve learning (24–26). However, the mentoring was informal, depended on the registrar’s personal style. Making mentorship part of formal training could help improve consistency. The Six – Part Model, under talent transformation calls for the extensive of feedback, coaching, and developmental assignments (8). To achieve this, there is need for the system to embrace a culture of assessment, recognition of challenges, and enhancement of support system in order to nurture the talents within organizations (27,28). The study also showed that registrars demonstrated ethical leadership in clinical practices. They handled difficult situations, such as when patients refused treatment because of personal beliefs or when patients could not afford medication. These situations require leaders to respect patient rights while giving the best care possible (29,30). For example, ethical principles demand that healthcare workers should act for the benefit of the patience (beneficence), and that healthcare workers have an obligation not to harm a patient (nonmalefidence), and acknowledgement that all person have got and should have the ability and power to make decisions and choices, and hence allowed for self-determination (autonomy), and an obligation to provide all the needed information regarding the provision of treatment and care for the patients’ decision making (informed consent) (31). Good ethical reasoning and analysis are skills that are essential to the patient management as healthcare workers encounter a broad spectrum of ethical dilemmas during clinical practices (32). Supervisees also noted that registrars were adaptable. They were able to manage busy ward conditions and respond to different learning needs. However, unclear role boundaries, especially between registrars and senior doctors sometimes led to confusion. Research shows that poor role clarity reduces teamwork and leadership confidence (26,27). Better communication, clear expectations, and structured orientation could help reduce confusion. Under the concept of capacity for complexity, innovation, and change, the Six-Part Model emphasizes the need for organizational fluidity such as the ability to influence, monitor, and respond to emerging changes such as shifts in workforce demographics, evolving patient needs, and technological advancements (7). However, the findings showed that participants were unsure whether the system was truly open to new ideas. Many displayed a limited willingness to learn and share new clinical practices, largely due to bureaucratic barriers that restrict innovation. Strengthening the system’s capacity for innovation will therefore require reducing these bureaucratic constraints and fostering an environment where learning, knowledge sharing, and the adoption of new clinical practices are encouraged and supported (33).Overall, the study shows that registrars already have important leadership strengths, especially in teamwork, mentorship, and ethical practice. But they need more support in system-level leadership, role clarity, formal mentorship, innovation, communication, and structured performance reviews. Improving these areas can help registrars become stronger leaders and improve the learning environment and patient care.

### Study limitations

The findings of this study should be interpreted in light of several limitations. First, the information provided by participants was largely based on their personal views and observations, which may not fully capture the breadth of experiences within the health system. Employing a mixed-methods approach such as triangulating focus group discussions with direct observations or document analysis could have strengthened the robustness of the data and reduced reliance on self-reported perspectives. Secondly, the investigators’ leadership experiences may have inadvertently introduced bias, particularly in data interpretation. Nevertheless, this risk was mitigated through peer debriefing and member checking. Finally, given the qualitative nature of the study, the findings are not statistically generalizable beyond the study setting. However, the insights generated are likely transferable to similar health system contexts and may inform policy and practice in comparable settings.

### Recommendations

To strengthen leadership, structured orientation and clear role definitions are recommended to reduce ambiguity and build confidence. Formal mentorship and feedback systems should be integrated to ensure consistent supervisee support, while enhanced communication, recognition, and relational leadership can foster trust and teamwork. Registrars’ participation in cross-departmental decision-making and resource mobilization should be encouraged, and organizational strategies that promote innovation and psychological safety can support experimentation and learning.

Based on these findings, the existing leadership training program could be enhanced by incorporating modules on role clarity, system-level leadership, structured mentorship and innovation management. Such improvements would equip registrars to navigate complex clinical and organizational environments, strengthening their leadership capacity, supervisee development, and patient care outcomes.

## Conclusion

Family Medicine registrars play a crucial role in clinical leadership, demonstrating strengths in collaboration, mentorship, ethical decision-making, resource stewardship, and adaptability. However, their effectiveness is limited by unclear role boundaries, informal mentorship, minimal system-level involvement, and insufficient structured feedback. Organizational culture and support mechanisms further constrain opportunities for innovation, relational leadership, and professional growth.

## Author contributions

**Conceptualisation:** Tony majo

**Formal analysis:** Tony Majo and Duncane Kwaitana

**Investigation:** Tony Majo and Martha Makwero

**Supervision:** Martha Makwero

**Writing – original draft:**Tony Majo and Duncane Kwaitana

**Writing – review & editing:** Tony Majo, Martha Makwero, Franchise Mambo and Duncane Kwaitana

## Data Availability

The data that support the findings of this study are available on request from the corresponding author (T.M.). The data are not publicly available because of privacy restrictions. Specifically, transcripts containing information (names and locations) that could compromise the privacy of research participants.

## Acknowledgement

The authors acknowledge the input of the participants and research assistants.

## References

1. World Health Organization. (2018). Declaration of Astana: Global Conference on Primary Health Care, Astana, Kazakhstan, 25 and 26 October 2018. World Health Organization. Retrieved from https://www.who.int/publications/i/item/WHO-HIS-SDS-2018.61

2. World Health Organization. (1978). Declaration of Alma-Ata: International Conference on Primary Health Care, Alma-Ata, USSR, 6–12 September 1978. World Health Organization. Retrieved from https://www.who.int/publications-detail-redirect/declaration-of-alma-ata

3. Johnson, O., et al. (2020). Interventions to strengthen the leadership capabilities of healthcare managers in sub-Saharan Africa. BMJ Global Health, 5(11), e003518. 10.1136/bmjgh-2020-003518

4. Kimball, A. M., et al. (2019). Strengthening public health leadership in Africa: An innovative fellowship program. Academic Medicine, 94(8), 1146–1149. 10.1097/ACM.0000000000002707

5. Manabe, Y. C., et al. (2020). Leadership training to accelerate progress in public health in sub-Saharan Africa: Time for action. The Lancet Global Health, 8(10), e1253–e1254. 10.1016/S2214-109X(20)30321-1

6. Government of the Republic of Malawi . (2022). Malawi’s Health Sector Strategic Plan III (HSSP III) for 2023–2030. Retrieved from https://faolex.fao.org/docs/pdf/mlw220703.pdf

7. Browning HW, Torain DJ, Patterson TE. *Collaborative healthcare leadership: A six-part model for adapting and thriving during a time of transformative change.* Greensboro (NC): Center for Creative Leadership; 2011

8. Peters M, ten Cate O. Bedside teaching in medical education: a literature review. Perspect Med Educ. 2014;3(2):76–88. doi:10.1007/s40037-013-0083-y

9. Sandelowski M. Sample size in qualitative research. Res Nurs Health. 1995;18(2):179–183. doi:10.1002/nur.4770180211.

10. Malterud K, Siersma VD, Guassora AD. Sample size in qualitative interview studies: guided by information power. Qual Health Res. 2016;26(13):1753–60.

11. Braun V, Clarke V. Using thematic analysis in psychology. Qual Res Psychol. 2006;3(2):77–101. doi: 10.1191/1478088706qp063oa

12. Lincoln YS, Guba EG. Naturalistic Inquiry. Newbury Park, CA: Sage Publications; 1985. doi: 10.1016/0147-1767(85)90062-8

13. Swanwick T, McKimm J. Clinical leadership development in postgraduate medical education. Br J Hosp Med. 2011;72(11):643–647.

14. Harden RM, Lilley P. The training of young clinicians in leadership. Med Teach. 2018;40(2):120–126.

15. Salas E, Sims DE, Burke CS. Is there a “Big Five” in teamwork? Small Group Res. 2005;36(5):555–599.

16. Reeves S, Lewin S, Espin S, Zwarenstein M. Interprofessional teamwork in health care. Wiley-Blackwell; 2011.

17. West MA, Lyubovnikova J. Illusions of team working in health care. J Health Organ Manag. 2013;27(1):134–142.

18. Frich JC, Brewster AL, Cherlin EJ, Bradley EH. Leadership development in health care: a systematic review. Acad Med. 2015;90(8):1128–1141.

19. Nadler DA, Tushman ML. The organization of the future. Organ Dyn. 1999;28(1):45–60.

20. Bowie C, Purcell R, Shava J. Managing health resources in low-income countries. Trop Med Int Health. 2019;24(2):123–130.

21. Daire J, Gilson L, Cleary S. Leadership and governance in the [Information redacted to maintain the integrity of the review process] an health system. Health Policy Plan. 2014;29 Suppl 2:ii59–ii69.

22. Mahmood K, Khan A. Accountability and efficiency in health care delivery. Int J Health Policy Manag. 2020;9(5):199–210.

23. Ulrich D, Allen J, Brockbank W, Younger J, Nyman M. HR Transformation. McGraw-Hill; 2009.

24. Kilminster S, Cottrell D, Grant J, Jolly B. Effective educational and clinical supervision. Med Teach. 2007;29(1):2–19.

25. Sambunjak D, Straus SE, Marušić A. Mentoring in academic medicine. JAMA. 2006;296(9):1103–1115.

26. Edmondson AC. The fearless organization. Wiley; 2019.

27. Eva KW, Bordage G, Campbell C, et al. Towards a program of assessment for health professionals. Med Teach. 2016;38(7):657–662.

28. Hawkins R, Epstein R. ACGME competencies: practice-based learning and improvement. J Grad Med Educ. 2012;4(2):193–198.

29. Beauchamp TL, Childress JF. Principles of biomedical ethics. 8th ed. Oxford University Press; 2019.

30. Gillon R. Medical ethics: four principles plus attention to scope. BMJ. 1994;309:184– 188.

31. Johnstone M-J. Bioethics: a nursing perspective. 7th ed. Elsevier; 2024.

32. Grady C. Ethical challenges in clinical practice. N Engl J Med. 2013;368:1742–1747.

33. Greenhalgh T, Papoutsi C. Studying complexity in health services research. BMC Med. 2018;16(1):95

